# Incidence and Predictors of In-stent Restenosis Following Intervention for Pulmonary Vein Stenosis due to Fibrosing Mediastinitis

**DOI:** 10.1101/2023.11.02.23298011

**Authors:** Mengfei Jia, Hongling Su, Kaiyu Jiang, Aqian Wang, Zhaoxia Guo, Hai Zhu, Fu Zhang, Pan Xin, Yunshan Cao

## Abstract

**Background:** Percutaneous transluminal pulmonary venoplasty (PTPV) is an emerging treatment for pulmonary vein stenosis (PVS) caused by fibrosing mediastinitis (FM). However, the incidence and predictors of in-stent restenosis (ISR) are elusive. We sought to identify the predictors of ISR in patients with PVS caused by extraluminal compression due to FM.

**Methods:** We retrospectively enrolled patients with PVS-FM who underwent PTPV between July 1, 2018, and December 31, 2022. According to ISR status, patients were divided into two groups: the ISR group and the non-ISR group. Baseline characteristics (demographics and lesions) and procedure-related information were abstracted from patient records and analyzed. Univariate and multivariate analyses were performed to determine the predictors of ISR.

**Results:** A total of 142 stents were implanted in 134 PVs of 65 patients with PVS-FM. Over a median follow-up of 6.6 (3.4-15.7) months, 61 of 134 PVs suffered from ISR. Multivariate analysis demonstrated a significantly lower risk of ISR in PVs with a larger reference vessel diameter (RVD) (odds ratio (OR): 0.79; 95% confidence interval [CI]: 0.64 to 0.98; *P*=0.032), and stenosis of the corresponding pulmonary artery (Cor-PA) independently increased the risk of restenosis (OR: 3.41; 95% CI: 1.31 to 8.86; *P*=0.012). The cumulative ISR was 6.3%, 21.4%, and 39.2% at the 3-, 6-, and 12-month follow-ups, respectively.

**Conclusion:** ISR is very high in PVS-FM, which is independently associated with RVD and Cor-PA stenosis.

Central illustration
Based on the constructed prediction model, the RVD and stenosis of Cor-PA were found to be independently associated with ISR, and their sensitivity and optimal cutoff values for the prediction of restenosis are shown in (a) and (b), respectively. The risk of ISR significantly increased when PA stenosis occurred; the risk of restenosis decreased significantly when the RVD was larger than 8.4 mm.

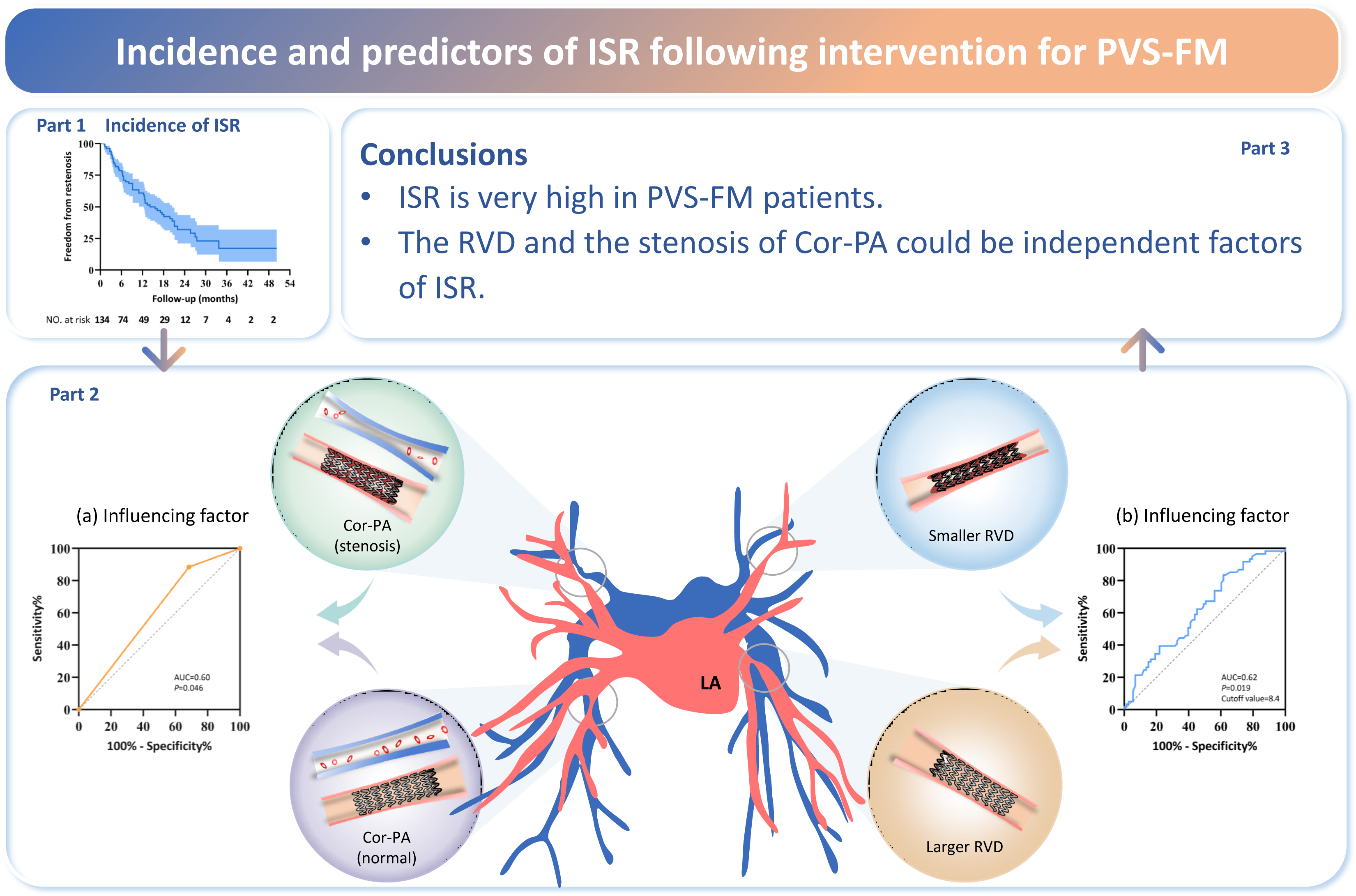

## Introduction

Fibrosing mediastinitis (FM) is characterized by benign proliferative fibrous tissue in the mediastinum, often compressing the pulmonary artery (PA), pulmonary vein (PV), bronchi, and superior vena cava, presenting with cough, dyspnea, hemoptysis, pleural effusion, superior vena cava syndrome, pulmonary hypertension, and right heart failure.^[1]^ Pulmonary vein stenosis (PVS) caused by FM (PVS-FM) is a kind of typical extraluminal compressing stenosis that is rare but fatal. Percutaneous transluminal pulmonary venoplasty (PTPV) is an emerging alternative for PVS-FM.^[2–4]^

The first balloon angioplasty (BA) reported by Massumi et al^[5]^ in 1981 was performed in a female patient with PVS-FM. However, early reports showed that BA was unsuccessful in the treatment of PVS, including a modified BA technique.^[6,7]^ The first report on stent venoplasty of PVS-FM was in 2001, which brought a new therapeutic modality for FM.^[8]^ Reviewing the progress of PV interventions, in the early stage, transcatheter angioplasty was mainly used to correct congenital or postoperative PVS in children.^[9]^ Since the report of PVS after pulmonary vein isolation (PVI) in 1998, catheter-based intervention has become increasingly common in the treatment of PVS caused by PVI (PVS-PVI) during the next 20 years.^[10]^ Nevertheless, detailed information is scarce about interventional treatment for PVS-FM, including hemodynamic changes, procedure-related complications, comprehensive follow-up data, incidence, and predictors of in-stent restenosis (ISR).^[11]^

The pathogenesis of PVS-FM is different from that of PVS-PVI and congenital PVS. PVS-FM is attributed to extraluminal proliferative fibrous tissue compression,^[12]^ while other PVS are attributed to intimal hyperplasia.^[13,14]^ Hence, even though PTPV has been successfully used in PVS-PVI, PTPV in PVS-FM might be different. Our preliminary data showed that patients with PVS-FM who underwent interventions, both in terms of hemodynamics and exercise capacity, had clinical improvement but a high prevalence of restenosis during a very short-term follow-up.^[15]^ Therefore, identifying the clinical and procedural factors associated with ISR is critical to guide intervention and optimize postintervention surveillance strategies. Against this background, we sought to identify the predictors associated with ISR following PVS-FM intervention.

## Methods

### Study population

From July 1, 2018, to December 31, 2022, we identified 144 patients with FM according to history, symptoms, signs, and findings in enhanced computed tomography (CT) with contrast in our center. Patients with PVS caused by tumors and other diseases were excluded. Patients were followed from PVS-FM diagnosis until the last follow-up or until May 2023 if receiving ongoing care. Patients were followed at set intervals with repeat CT imaging at 1-3, 3-6, 6-12, and 24 months after the intervention.

### Data collection

Patient clinical data at baseline and follow-up periods were collected. If a patient with PVS-FM was analyzed as part of the ISR cohort and the patient also had normal veins, the patient might have had veins analyzed in 2 different cohorts. The procedure-related parameters collected included minimal lumen diameter (MLD), lesion length, and reference vessel diameter (RVD) (taken as the mean diameter of the normal-appearing proximal and distal segment; if the PV diameter at both ends of the stenotic site was greatly different, the diameter of the distal PV served as the reference diameter). Furthermore, the maximal balloon diameter (using the actual measured maximal balloon size), maximal balloon inflation pressure, stent diameter, stent length, maximal stent inflation pressure, final lumen diameter (FLD), balloon-to-vessel ratio (calculated as the largest diameter of the inflated balloon divided by RVD), vessel-to-stent ratio (calculated as the FLD divided by stent diameter), and diameter stenosis (%) (calculated as [1-(MLD/RVD)]×100%) were also included. In addition, PA narrowing in series with stenotic PV, pleural effusion, and postoperative anticoagulants was collected. The pulmonary venous flow grade (PVFG) was assigned using grades 0-3.^[15]^

### Outcomes

The primary endpoint of the study was the incidence of ISR following PTPV or pulmonary angiography during the follow-up period, and the secondary endpoints were the World Health Organization functional class (WHO-FC) and 6-minute walking distance (6MWD). We also analyzed the demographic, clinical and procedural variables associated with ISR.

Second, we sought to understand the outcomes of patients with PVS-FM who underwent pulmonary vein intervention only.

ISR was defined as stenosis >50% of the vessel size as confirmed by repeated angiography or an increase in pressure gradient (Pd) (≥5 mmHg) across the stenotic site compared to the last measurement.

### Research ethics and patient consent

The ethics committee of Gansu Provincial Hospital reviewed and approved the study protocol (2022-302) on 25, August 2022 as well as granted exemption from obtaining informed consent from patients.

### Data availability

Anonymized data used and/or analyzed during the current study are available from the corresponding author upon reasonable request from any qualified investigator for the sole purpose of the study.

### Statistical analysis

Categorical data are expressed as counts and proportions (%). Continuous data are reported as the mean ± SD or as the median (interquartile range). The Kolmogorov‒ Smirnov test was used to determine the normality of the data distribution. For continuous variables, *t* tests or Wilcoxon rank sum tests were used as appropriate. For categorical variables, χ^2^ tests or Fisher exact tests were used. A binary logistic analysis was used to construct an optimal model in multiple variables analysis. Receiver operating characteristic (ROC) analyses were used to determine the predictive power of variables for ISR. A 95% confidence interval (CI) is provided for all estimates. A *P* value<0.05 was considered significant. The Kaplan‒Meier method was used to estimate and plot the time curves for the appearance of restenosis in the initial intervention vessels, and the log-rank test was used to compare restenosis between the different sizes of RVD. Statistical analysis was performed using SPSS version 25.0 (*SPSS, Chicago, Illinois*), and figures were plotted by GraphPad Prism software v.8.0 (*GraphPad Software, San Diego, California USA*).

## Results

### Baseline characteristics

The study flowchart is shown in **Figure 1**. Of 144 patients with FM during the study period, 43 patients who did not undergo PV intervention, 2 patients with unsuccessful PV intervention, and 7 patients who underwent balloon pulmonary venoplasty (BPV) alone were excluded. A total of 92 patients successfully underwent percutaneous transluminal stent venoplasty. Of them, twenty-five (27.2%) patients were lost to follow-up, 2 (2.2%) had no imaging data at follow-up, and 65 patients (70.7%) with 142 stents implanted in 134 lesioned PVs during 72 sessions underwent CT and/or selective pulmonary venographic surveillance at a median of 6.6 (3.4-15.7) months of follow-up. Of the 65 patients ultimately included in the analysis, 2 veins were implanted with stents directly without ballooning, 8 veins were implanted with 2 overlapping stents due to long-segment lesions, and 126 veins were stented after initial balloon venoplasty failed to improve the Pd across the stenotic site. The baseline characteristics are shown in **Table 1**, and the procedural and lesion characteristics are shown in **Table 2**.

**Figure 1.**
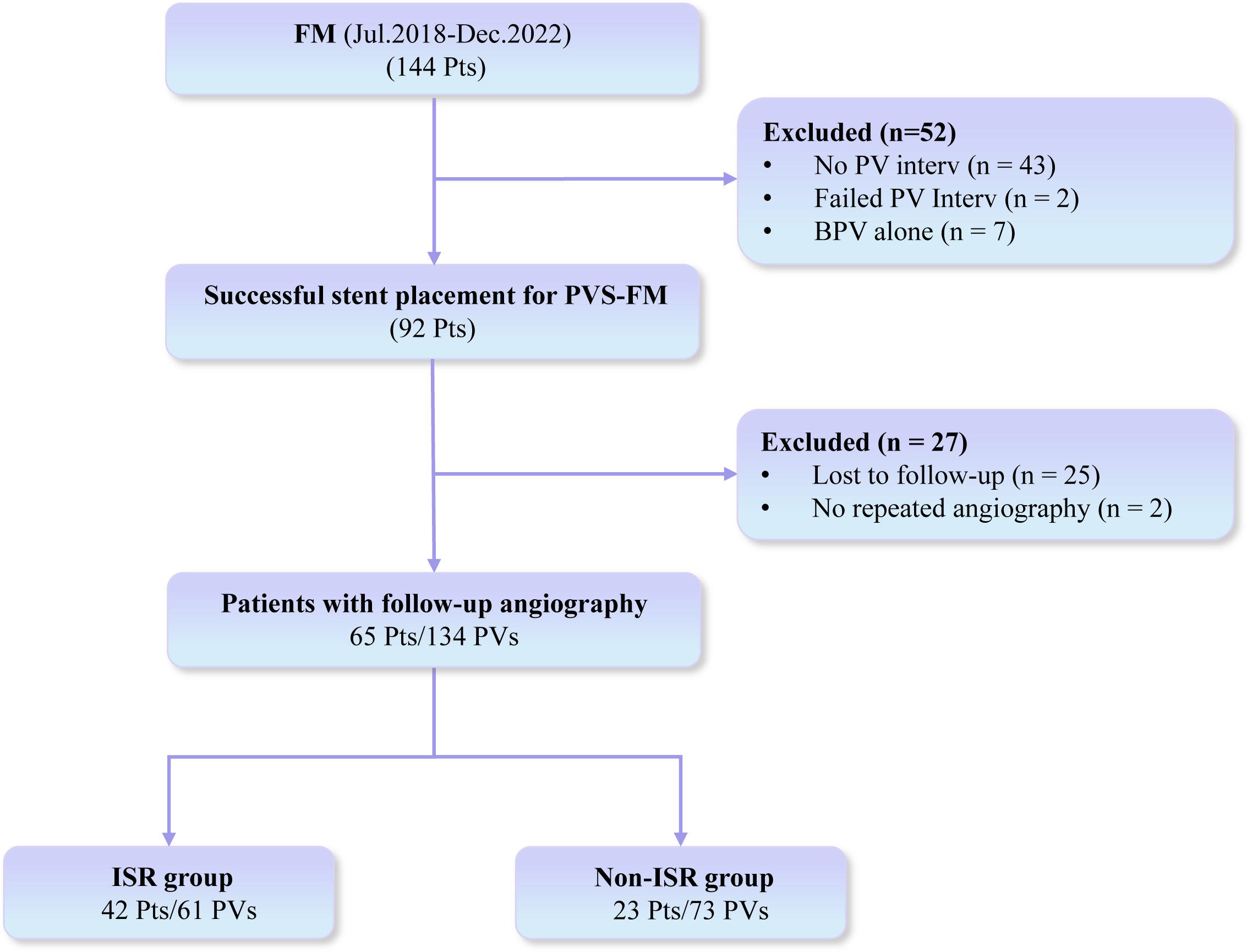
Flowchart of patient enrollment.

**Table 1.** Baseline characteristics.

|  | **Total (N=65)** | **ISR (n=42)** | **Non-ISR (n=23)** | **P Value** |
| --- | --- | --- | --- | --- |
| *Demographics* |  |  |  |  |
| Age, years | 66.0 (60.0, 70.0) | 64.5 (59.8, 69.3) | 67.0 (60.0, 72.0) | 0.38 |
| Body mass index, kg/m<sup>2</sup> (N=64) | 22.4 ± 3.6 | 22.2 ± 3.5 | 22.7 ± 3.7 | 0.53 |
| Female | 29 (44.6) | 20 (47.6) | 9 (39.1) | 0.51 |
| Duration of symptoms, months | 37.0 (24.0, 72.0) | 38.0 (24.0, 72.0) | 36.0 (14.0, 96.0) | 0.83 |
| *Clinical presentation* |  |  |  |  |
| Dyspnea | 61 (93.8) | 38 (90.5) | 23 (100.0) | 0.29 |
| Cough | 20 (30.8) | 15 (35.7) | 5 (21.7) | 0.24 |
| Hemoptysis | 4 (6.2) | 4 (9.5) | 0 (0.0) | 0.29 |
| Chest distress | 40 (61.5) | 22 (52.4) | 18 (78.3) | 0.040 |
| Palpitations | 3 (4.6) | 2 (4.8) | 1 (4.3) | 1.00 |
| Edema of lower limbs | 17 (26.2) | 12 (28.6) | 5 (21.7) | 0.55 |
| Pleural effusion | 49 (75.4) | 32 (76.2) | 17 (73.9) | 0.84 |
| *Comorbidity* |  |  |  |  |
| Hypertension | 21 (32.3) | 14 (33.3) | 7 (30.4) | 0.81 |
| Diabetes mellitus | 14 (21.5) | 7 (16.7) | 7 (30.4) | 0.22 |
| COPD | 48 (73.8) | 29 (69.0) | 19 (82.6) | 0.23 |
| Atelectasis | 42 (64.6) | 29 (69.0) | 13 (56.5) | 0.31 |
| Tuberculosis | 39 (60.0) | 26 (61.9) | 13 (56.5) | 0.67 |
| *Hemodynamics (n=64)* |  |  |  |  |
| SaO<sub>2</sub>, % | 89.0 (87.3, 92.0) | 89.0 (87.0, 92.0) (n=41) | 90.0 (88.0, 91.0) | 0.83 |
| mRAP, mmHg | 3.0 (2.0, 5.0) | 3.0 (2.0, 4.5) (n=41) | 5.0 (2.0, 7.0) | 0.067 |
| sPAP, mmHg | 70.5 (54.0, 80.8) | 71.0 (53.5, 81.0) (n=41) | 68.0 (54.0, 78.0) | 0.70 |
| dPAP, mmHg | 27.0 (22.0, 33.0) | 27.0 (22.0, 33.0) (n=41) | 27.0 (20.0, 34.0) | 0.80 |
| mPAP, mmHg | 39.5 (33.0, 50.0) | 40.0 (33.0, 49.5) (n=41) | 39.0 (30.0, 53.0) | 0.88 |
| PAWP, mmHg (n=63) | 7.0 (6.0, 9.0) | 7.0 (6.0, 9.0) (n=40) | 7.0 (6.0, 7.0) | 0.97 |
| PVR, WU | 6.3 (5.3, 9.9) | 6.3 (5.2, 9.6) (n=41) | 6.8 (5.5, 10.0) | 0.48 |
| CO, L/min | 4.6 (3.8, 5.4) | 4.8 (3.9, 5.5) (n=41) | 4.4 (3.7, 5.3) | 0.34 |
| CI, L/min/m<sup>2</sup> | 2.8 ± 0.6 | 3.0 ± 0.7 (n=41) | 2.8 ± 0.6 | 0.23 |
| SvO<sub>2</sub>, % | 62.9 ± 9.0 | 63.6 ± 9.1 (n=41) | 61.7 ± 9.0 | 0.52 |
| *Echocardiographic* |  |  |  |  |
| LA size, mm | 32.4 ± 5.0 | 32.4 ± 4.9 | 32.3 ± 5.2 | 0.94 |
| TAPSE, mm (n=50) | 19.2 ± 4.6 | 19.7 ± 4.0 (n=33) | 18.2 ± 5.5 (n=17) | 0.30 |
| RAA, end-systolic, cm<sup>2</sup> (n=42) | 16.5 (14.0, 20.6) | 15.8 (14.0, 22.5) (n=27) | 17.5 (14.3, 20.4) (n=15) | 0.66 |
| RVA, end-diastolic, cm<sup>2</sup> (n=42) | 24.4 ± 9.2 | 23.5 ± 9.0 (n=27) | 26.0 ± 9.7 (n=15) | 0.41 |
| *Exercise capacity* |  |  |  |  |
| WHO-FC, I/II/III/IV | 0/25/32/8 | 0/17/19/6 | 0/8/13/2 | 0.64 |
| 6MWD, m (n=42) | 306.4 ± 97.8 | 287.6 ± 100.1 (n=27) | 340.2 ± 86.6 (n=15) | 0.16 |
| *Laboratory values* |  |  |  |  |
| NT-proBNP, pg/ml | 732.6 (163.5, 1568.5) | 714.0 (128.3, 1374.3) | 732.6 (203.0, 2678.0) | 0.88 |
| NLR (n=62) | 4.7 (3.8, 6.8) | 4.6 (3.8, 5.8) (n=41) | 5.2 (3.3, 8.9) (n=21) | 0.48 |
| CRP, mg/L (n=59) | 5.8 (1.7, 14.1) | 6.3 (1.7, 16.9) (n=40) | 5.6 (1.7, 9.4) (n=19) | 0.75 |
| D-Dimer, ug/ml (n=58) | 1.0 (0.6, 1.5) | 1.0 (0.7, 1.5) (n=36) | 0.9 (0.3, 1.4) (n=22) | 0.22 |
| With PAS | 63 (96.9) | 41 (97.6) | 22 (95.7) | 1.00 |

Values are mean ± SD, n (%), or M (Q1, Q3). 6MWD = 6-minute walking distance; CI = cardiac index, CO = cardiac output; COPD = chronic obstructive pulmonary disease; CRP = C-reactive protein; dPAP = diastolic pulmonary artery pressure; ISR = in stent restenosis; LA = left atrial; mPAP = mean pulmonary artery pressure; NLR = Neutrophil-to-Lymphocyte ratio; NT-proBNP = N-terminal pro-brain natriuretic peptide; PAS = pulmonary artery stenosis; PAWP = pulmonary artery wedge pressure; PVR = pulmonary vascular resistance; RA = right atrial; mRAP = mean right atrial pressure; RV = right ventricle; SaO<sub>2</sub> = arterial oxygen saturation; sPAP = systolic pulmonary artery pressure; SvO<sub>2</sub> = mixed venous oxygen saturation; TAPSE = tricuspid annular plane systolic excursion; WHO-FC = World Health Organization functional class.

**Table 2.**
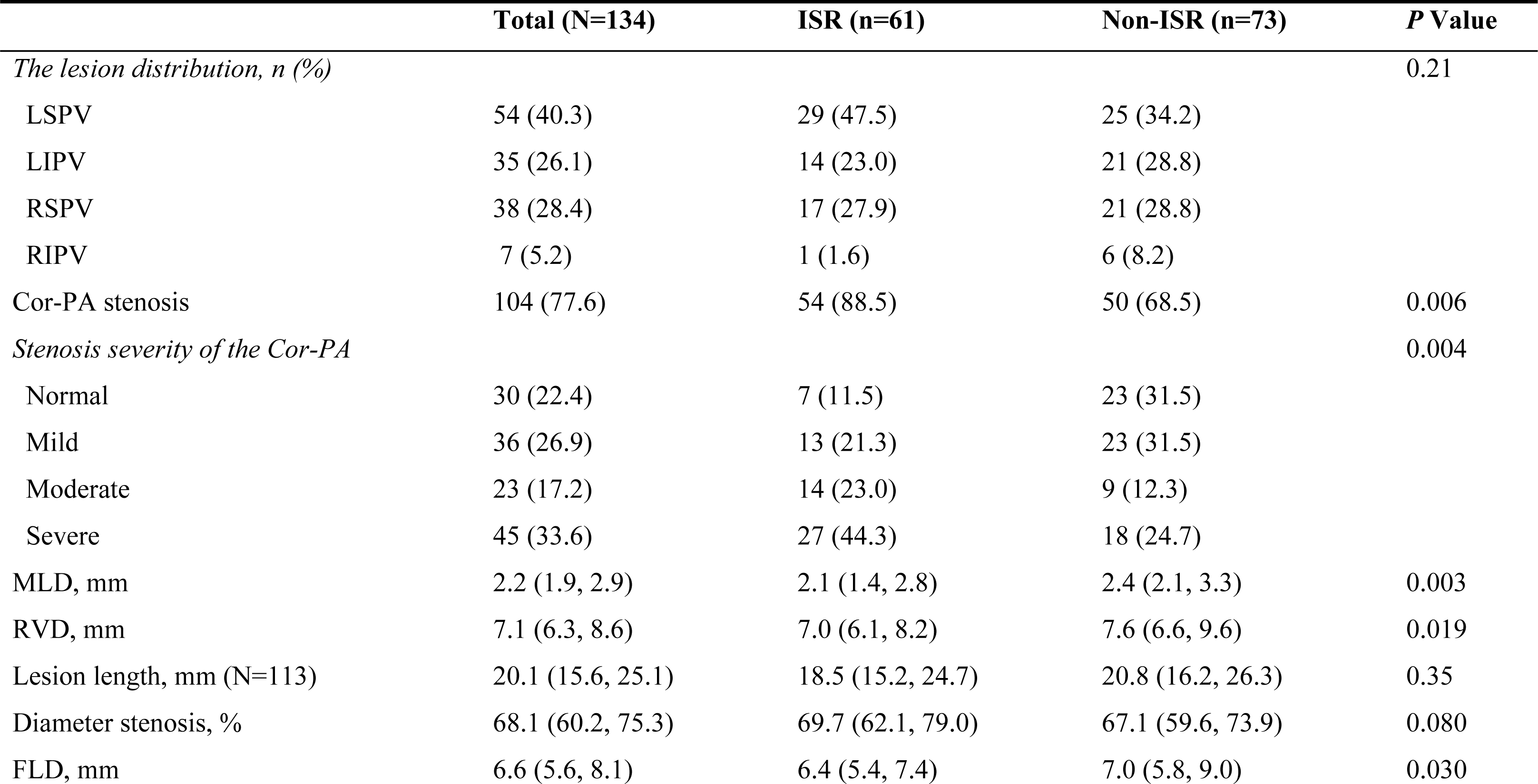
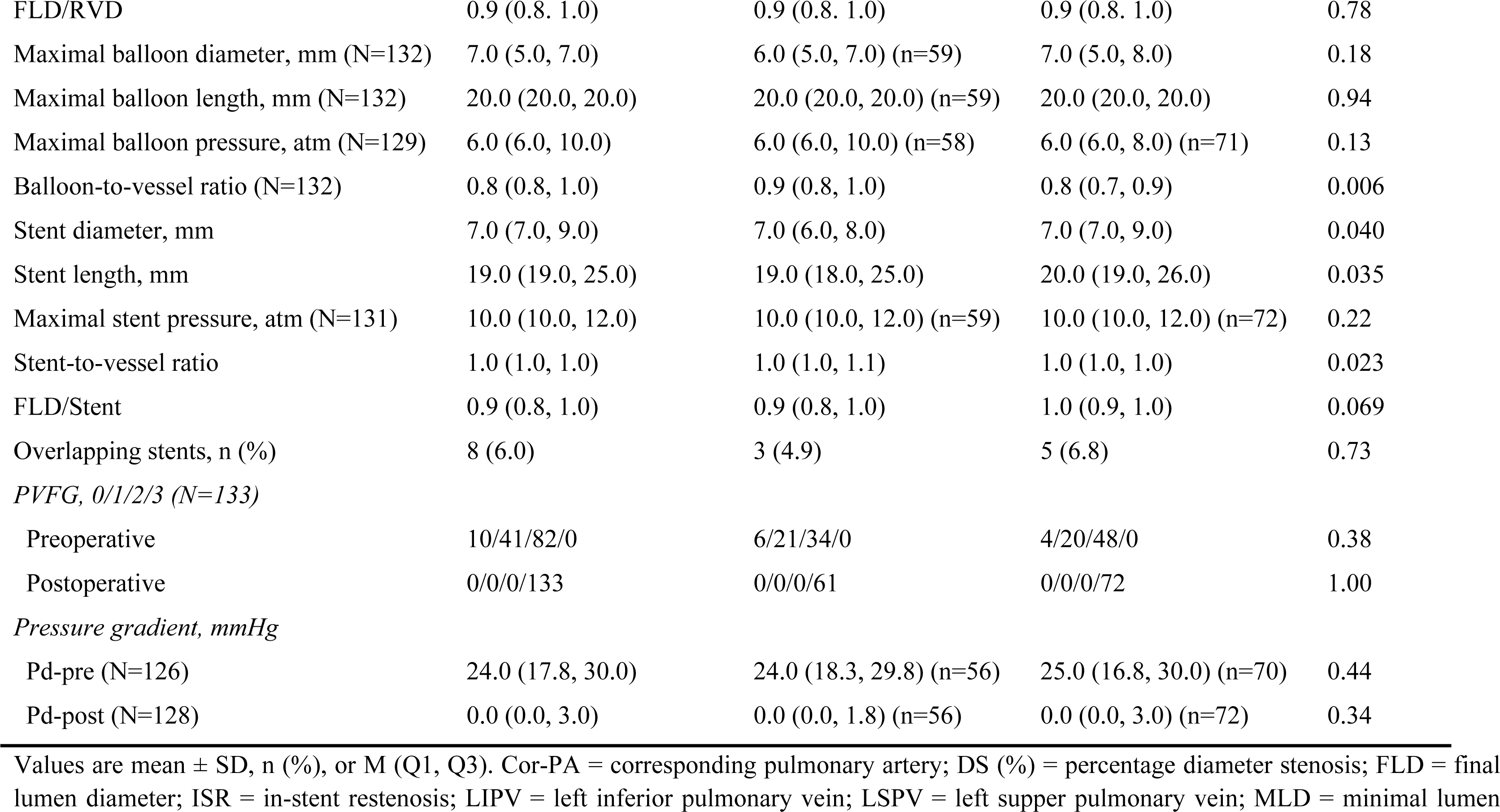
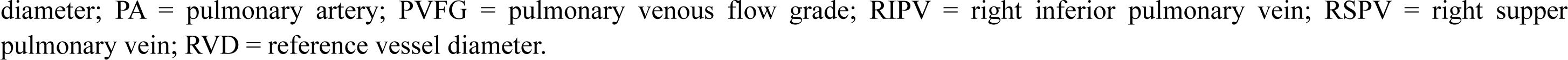
Lesion characteristics and procedural-related information.

### Incidence of in-stent restenosis (ISR)

At a median follow-up of 6.6 (3.4-15.7) months, ISR was found in 61 of 134 treated veins. The cumulative ISR was 6.3%, 21.4%, and 39.2% at the 3-, 6-, and 12-month follow-ups, respectively (**Figure 2**).

**Figure 2.**
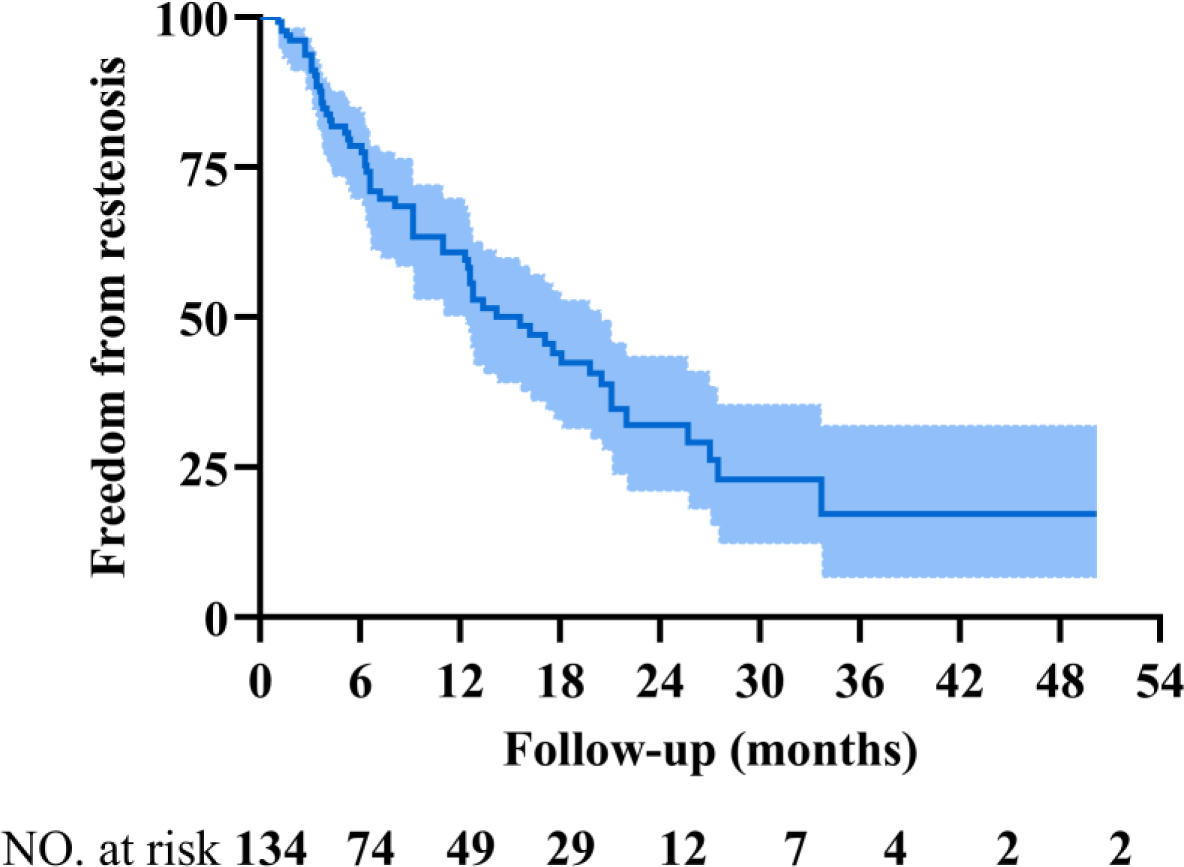
Cumulative incidence of ISR. Kaplan‒Meier curve depicting the probability of ISR over a median of 6.6 (3.4-15.7) months.

### Univariate analysis

Patients with and without ISR had similar ages, sex distribution, body mass index, and medical histories, including chronic obstructive pulmonary disease, diabetes mellitus (DM), tuberculosis, etc. There were also no significant differences in the hemodynamic and laboratory parameters between the two groups. The analysis of clinical factors failed to identify any factors significantly correlated with ISR. Among procedure-related factors, MLD, RVD, FLD, stent diameter, stent length, stent-to-vessel ratio, and stenosis of the corresponding pulmonary artery (Cor-PA) were associated with ISR (**Table 3**). **Multivariate analysis**

**Table 3.**
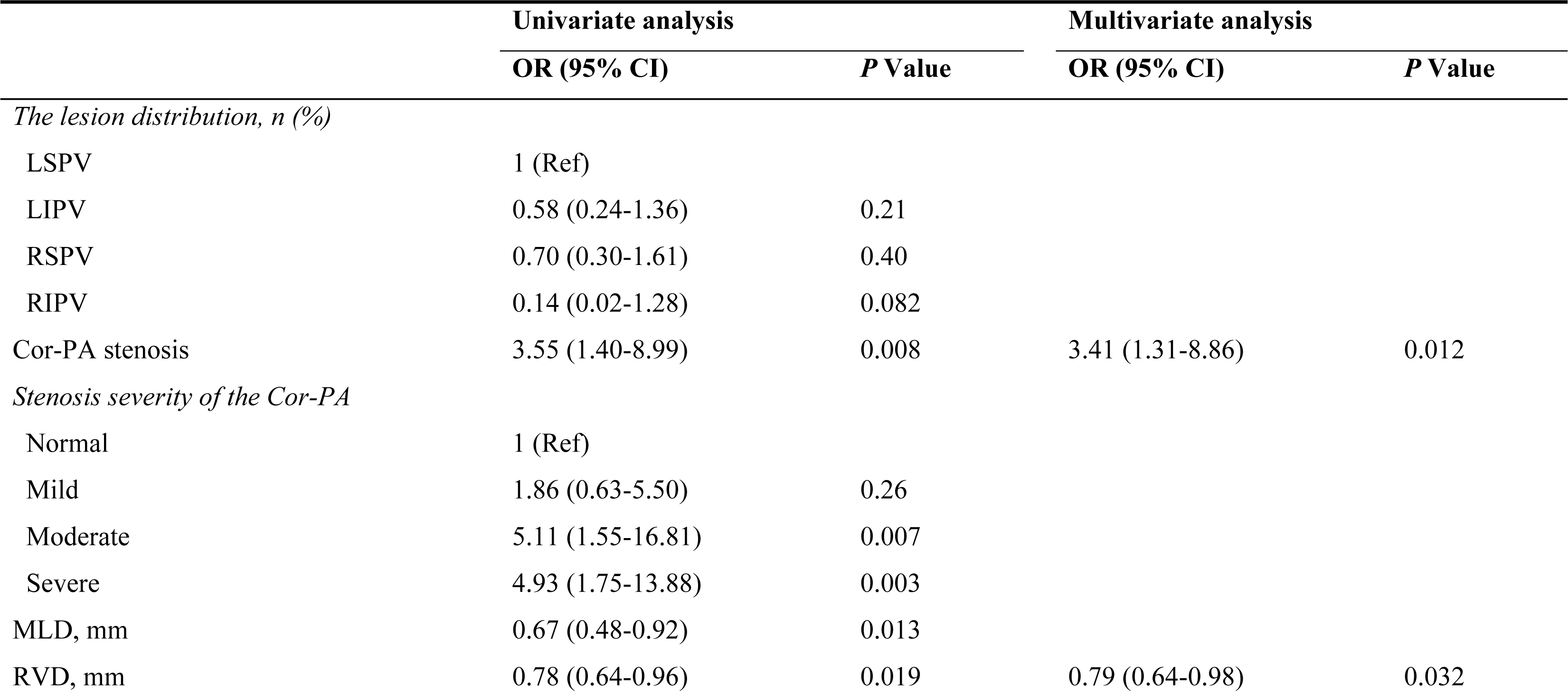
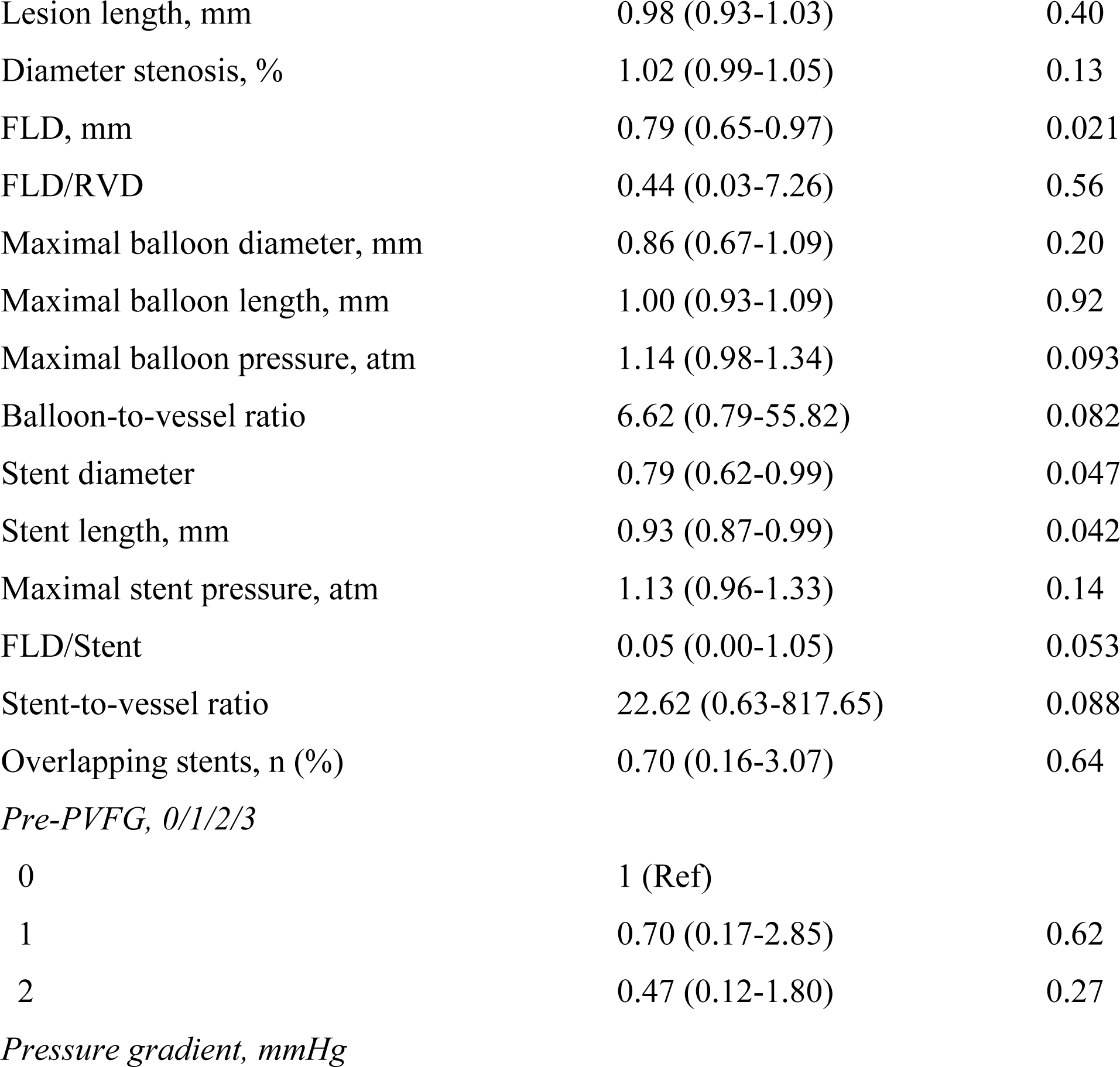
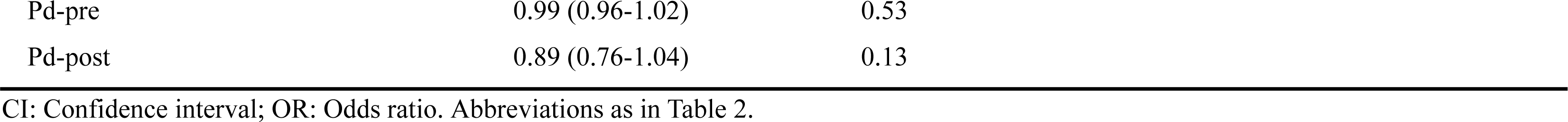
Per-vessel univariate and multivariate analysis associated with in-stent restenosis.

|  | Univariate analysis |  | Multivariate analysis |  |
| --- | --- | --- | --- | --- |
|  | OR (95% CI) | P Value | OR (95% CI) | P Value |
| *The lesion distribution, n (%)* |  |  |  |  |
| LSPV | 1 (Ref) |  |  |  |
| LIPV | 0.58 (0.24-1.36) | 0.21 |  |  |
| RSPV | 0.70 (0.30-1.61) | 0.40 |  |  |
| RIPV | 0.14 (0.02-1.28) | 0.082 |  |  |
| Cor-PA stenosis | 3.55 (1.40-8.99) | 0.008 | 3.41 (1.31-8.86) | 0.012 |
| *Stenosis severity of the Cor-PA* |  |  |  |  |
| Normal | 1 (Ref) |  |  |  |
| Mild | 1.86 (0.63-5.50) | 0.26 |  |  |
| Moderate | 5.11 (1.55-16.81) | 0.007 |  |  |
| Severe | 4.93 (1.75-13.88) | 0.003 |  |  |
| MLD, mm | 0.67 (0.48-0.92) | 0.013 |  |  |
| RVD, mm | 0.78 (0.64-0.96) | 0.019 | 0.79 (0.64-0.98) | 0.032 |

|  |  |  |
| --- | --- | --- |
| Lesion length, mm | 0.98 (0.93-1.03) | 0.40 |
| Diameter stenosis, % | 1.02 (0.99-1.05) | 0.13 |
| FLD, mm | 0.79 (0.65-0.97) | 0.021 |
| FLD/RVD | 0.44 (0.03-7.26) | 0.56 |
| Maximal balloon diameter, mm | 0.86 (0.67-1.09) | 0.20 |
| Maximal balloon length, mm | 1.00 (0.93-1.09) | 0.92 |
| Maximal balloon pressure, atm | 1.14 (0.98-1.34) | 0.093 |
| Balloon-to-vessel ratio | 6.62 (0.79-55.82) | 0.082 |
| Stent diameter | 0.79 (0.62-0.99) | 0.047 |
| Stent length, mm | 0.93 (0.87-0.99) | 0.042 |
| Maximal stent pressure, atm | 1.13 (0.96-1.33) | 0.14 |
| FLD/Stent | 0.05 (0.00-1.05) | 0.053 |
| Stent-to-vessel ratio | 22.62 (0.63-817.65) | 0.088 |
| Overlapping stents, n (%) | 0.70 (0.16-3.07) | 0.64 |
| *Pre-PVFG, 0/1/2/3* |  |  |
| 0 | 1 (Ref) |  |
| 1 | 0.70 (0.17-2.85) | 0.62 |
| 2 | 0.47 (0.12-1.80) | 0.27 |

*Pressure gradient, mmHg*
|  |  |  |
| --- | --- | --- |
| Pd-pre | 0.99 (0.96-1.02) | 0.53 |
| Pd-post | 0.89 (0.76-1.04) | 0.13 |

CI: Confidence interval; OR: Odds ratio. Abbreviations as in Table 2.

To guarantee that there was no multicollinearity among the variables, the appropriate variables were selected by calculating the tolerance and variance inflation factor (VIF). Then, a covariance diagnosis on the indicators that were significant in the results of univariate analysis and on the variables that might be clinically significant was performed. The variables with tolerances less than 0.1 and a VIF greater than 10 were excluded, while the remaining variables were analyzed through multivariate analysis. Procedure-related parameters that were independently associated with ISR included the RVD and the stenosis of Cor-PA **(central illustration)**. For ISR, RVD was associated with an adjusted OR of 0.79 (95% CI, 0.64 to 0.98, *P*=0.032), while the stenosis of Cor-PA was associated with an adjusted OR of 3.41 (95% CI, 1.31 to 8.86, *P*=0.012). The results of the ROC analysis for procedure-related variables for ISR are depicted in **Figure 3**. RVD>8.4 mm could be used as the cutoff point to predict ISR, and its sensitivity and specificity were 0.84 and 0.38, respectively. The subgroup of vessels with a reference diameter >8.4 mm had a significantly lower risk of ISR than the subgroup with a reference diameter ≤8.40 mm **(Figure 3D)**. Meanwhile, the sensitivity and specificity of Cor-PA stenosis were 0.89 and 0.69, respectively. When the positive and negative influencing factors were combined, the sensitivity and specificity were 0.77 and 0.55, respectively. Hence, we obtained the regression equation of related variables and restenosis: Logit (*P*)=0.614-0.236×RVD+1.226×Cor-PA stenosis (Cor-PA stenosis =1 if present, 0 absent).

**Figure 3.**
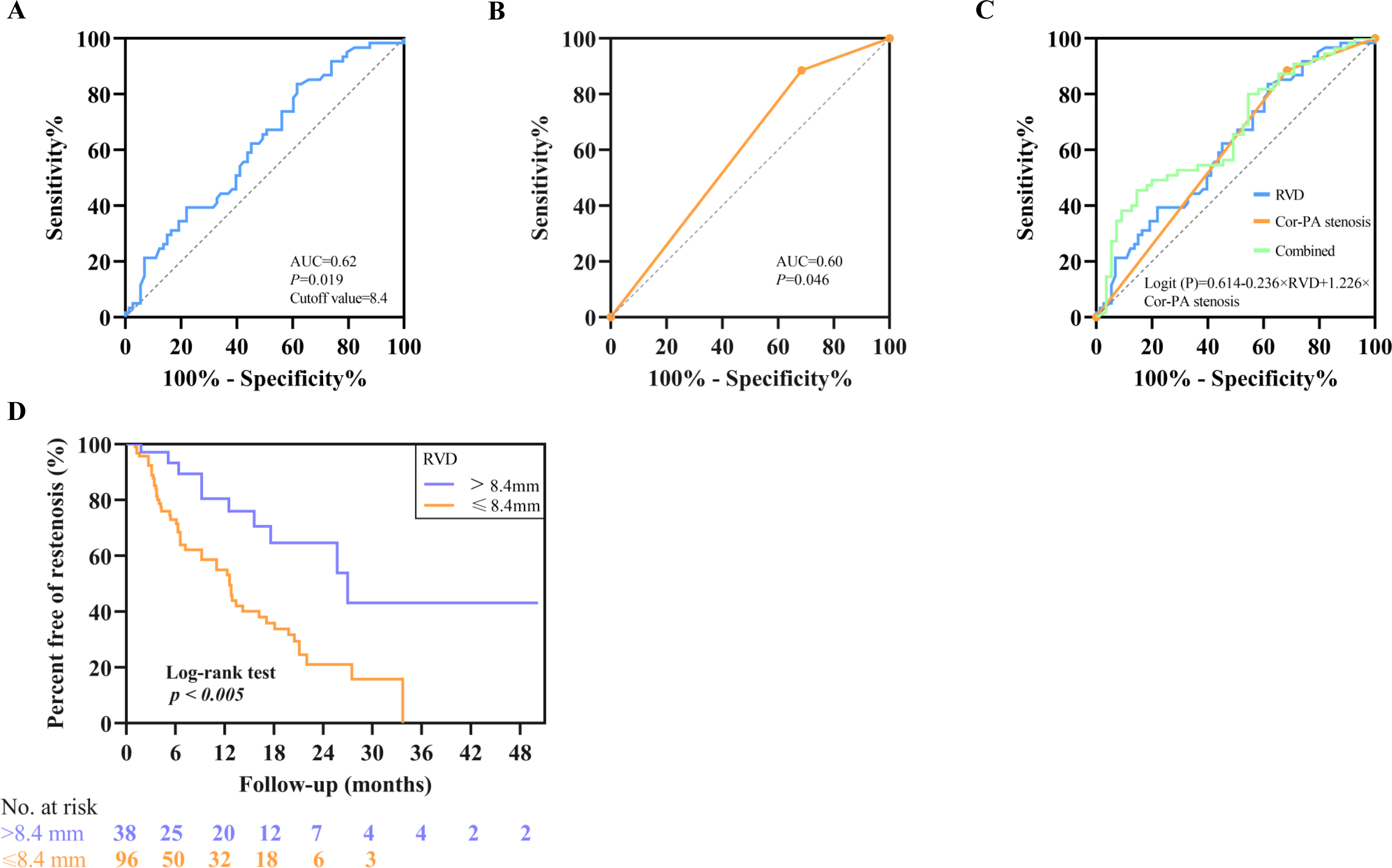
ROC analysis for the determination of ISR in the PVS-FM. (A) RVD had a sensitivity and specificity of 0.84 and 0.38, respectively, for a cutoff point of 8.4 mm (AUC, 0.62; 95% CI, 0.52 to 0.71; *P*=0.019) (blue line). (B) Cor-PA stenosis had a sensitivity and specificity of 0.89 and 0.69, respectively (AUC, 0.60; 95% CI, 0.51 to 0.70; *P*=0.046) (yellow line). (C) Binary logistic regression analysis rendered the following formula for the prediction of ISR: Logit (*P*)=0.614-0.236×RVD+1.226×Cor-PA stenosis (AUC, 0.68; 95% CI, 0.59 to 0.77; *P*<0.001) (green line). (D) Kaplan–Meier survival plot comparing freedom from restenosis stratified by RVD. There was a significant difference (*P*<0.005 for a log-rank test) between the RVD≤8.4 mm (orange solid line) and >8.4 mm (purple dashed line) groups.

### Procedural complications

In the analysis of 72 sessions performed in 65 patients, there were 13 episodes of chest tightness (18%) and 14 episodes of cough (19%), which were the most common during the intervention **(Supplemental Table 1)**. Mild hemoptysis and transient cardiac arrest/bradycardia occurred in 7% and 4% of sessions, respectively, with no requirement for additional intervention. There were 2 (3%) patients experiencing PV dissection/perforation who underwent balloon occlusion with low inflation pressure and recovered without any hemodynamic insults. One patient suffered from suspected acute pulmonary edema with acute onset of dyspnea and elevated left atrial pressure after stent implantation, high flow oxygen, and diuretics were administered, and these symptoms were relieved soon after. There were no cases of peri-procedure death or major hemoptysis occurred.

### Immediate and short-term efficacy

The MLD, Pd, and PVFG in the recruited patients were evaluated pre- and postintervention and drastically improved after the intervention. The MLD increased from 2.2 (1.9, 2.9) mm to 6.6 (5.6, 8.1) mm, and PVFG and Pd were significantly improved (*P*<0.001 for all) **(Figure 4A-C)**. Additionally, due to 17 patients undergoing PA intervention at the same time or later, the short-term efficacy of the remaining 43 (66.2%) patients with only PV intervention at follow-up was analyzed.

**Figure 4.**
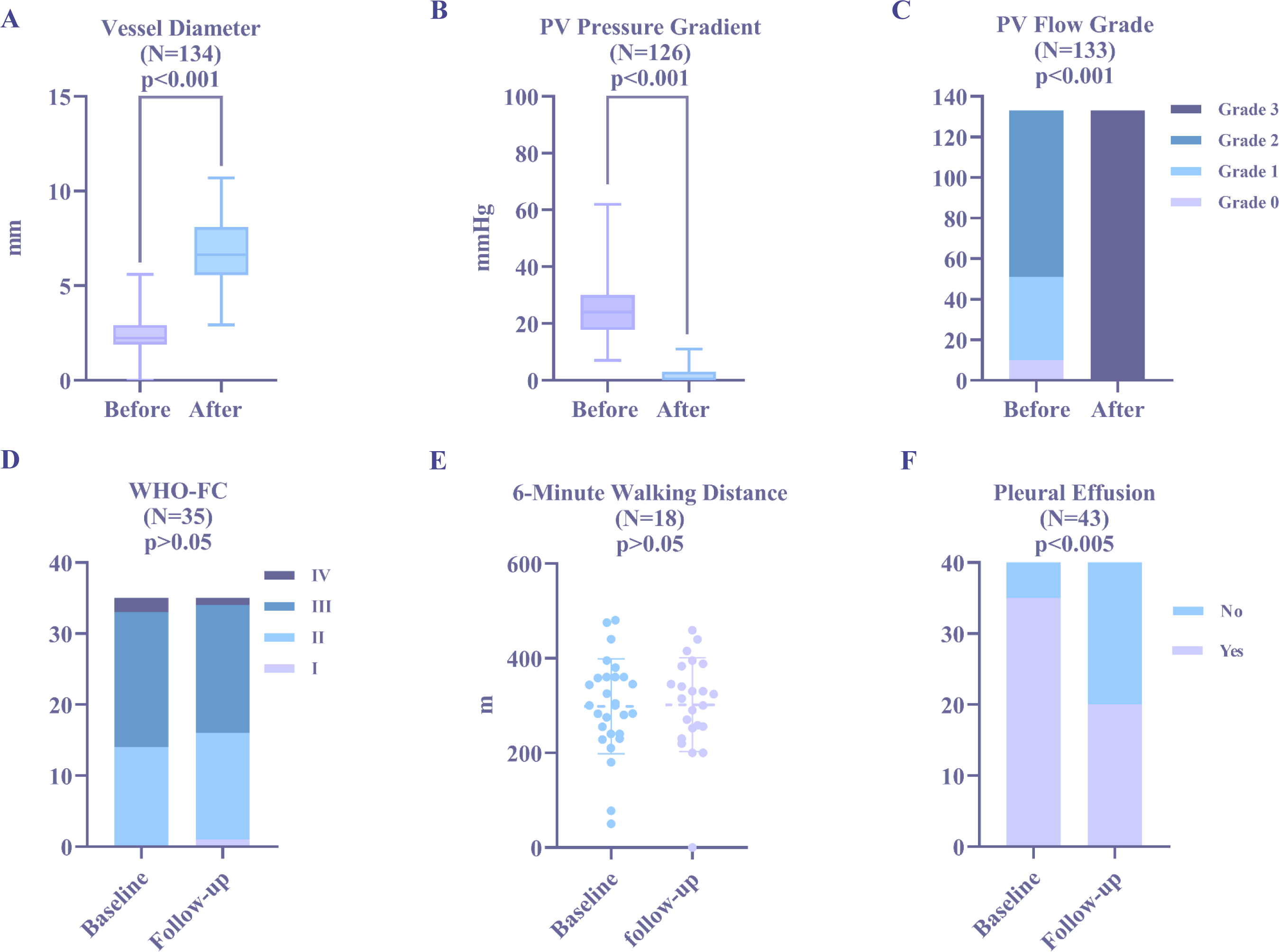
Immediate and short-term efficacy. A-C show the immediate effects of the intervention. D-F show the short-term effects of PV intervention alone. When the pre- and postintervention data were compared, there was a significant improvement in MLD, Pd, and PVFG (*P*<0.001 for all). There was a significant improvement in pleural effusion but no changes in WHO-FC and 6MWD after PV intervention compared with baseline (*P*<0.005, *P*>0.05, and *P*>0.05, respectively).

The median follow-up was 5.0 (3.1-11.2) months. Among the 43 patients, 23 underwent right heart catheterization during the follow-up. Comparisons of the baseline and follow-up data in patients with PV intervention are shown in **Table 4**. The pleural effusion decreased from 35 (81.3%) to 20 (46.5%) (3 of which were new pleural effusions) (*P*<0.005) during the follow-up. However, there was no improvement in the postoperative WHO-FC or 6MWD (*P*>0.05) **(Figure 4D-F)**. The mean PAP had a significant improvement (*P*=0.016), and there was an increase in left atrial size (*P*=0.439) but with no statistical significance.

**Table 4.**
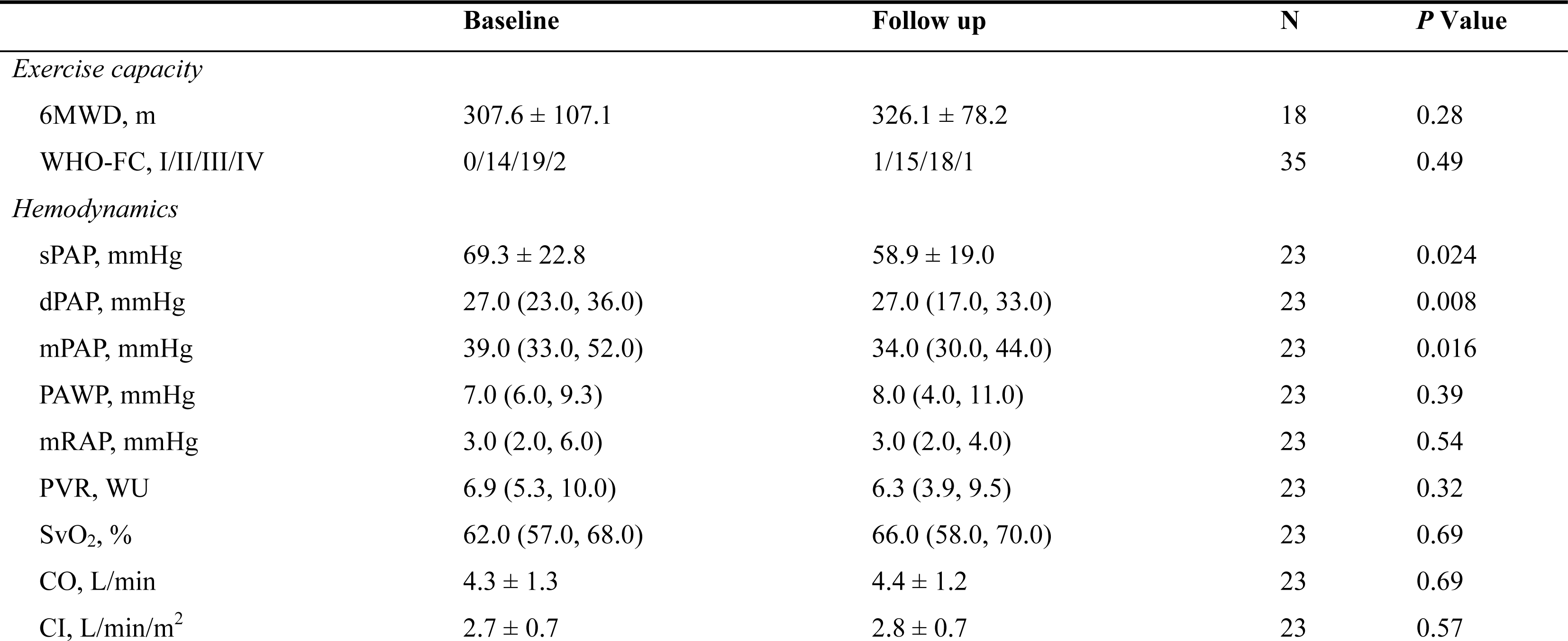
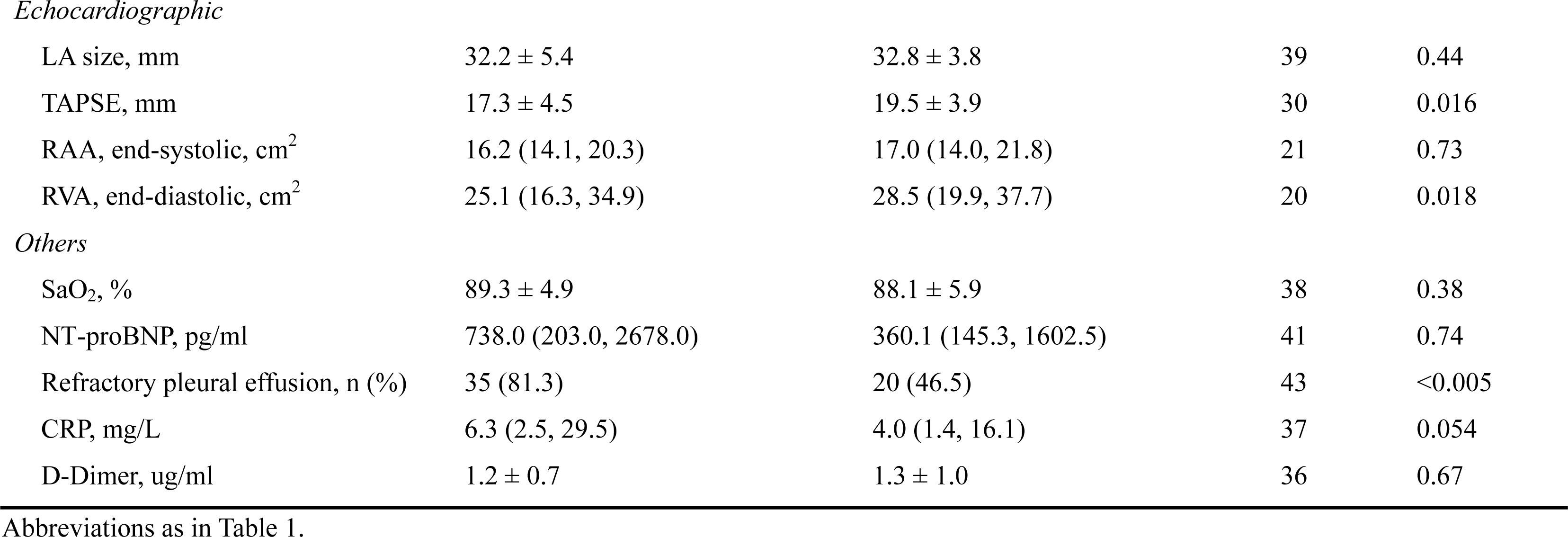
Short-term efficacy of percutaneous pulmonary venoplasty in patients with PVS-FM.

|  | Baseline | Follow up | N | P Value |
| --- | --- | --- | --- | --- |
| *Exercise capacity* |  |  |  |  |
| 6MWD, m | 307.6 ± 107.1 | 326.1 ± 78.2 | 18 | 0.28 |
| WHO-FC, I/II/III/IV | 0/14/19/2 | 1/15/18/1 | 35 | 0.49 |
| *Hemodynamics* |  |  |  |  |
| sPAP, mmHg | 69.3 ± 22.8 | 58.9 ± 19.0 | 23 | 0.024 |
| dPAP, mmHg | 27.0 (23.0, 36.0) | 27.0 (17.0, 33.0) | 23 | 0.008 |
| mPAP, mmHg | 39.0 (33.0, 52.0) | 34.0 (30.0, 44.0) | 23 | 0.016 |
| PAWP, mmHg | 7.0 (6.0, 9.3) | 8.0 (4.0, 11.0) | 23 | 0.39 |
| mRAP, mmHg | 3.0 (2.0, 6.0) | 3.0 (2.0, 4.0) | 23 | 0.54 |
| PVR, WU | 6.9 (5.3, 10.0) | 6.3 (3.9, 9.5) | 23 | 0.32 |
| SvO<sub>2</sub>, % | 62.0 (57.0, 68.0) | 66.0 (58.0, 70.0) | 23 | 0.69 |
| CO, L/min | 4.3 ± 1.3 | 4.4 ± 1.2 | 23 | 0.69 |
| CI, L/min/m<sup>2</sup> | 2.7 ± 0.7 | 2.8 ± 0.7 | 23 | 0.57 |
| *Echocardiographic* |  |  |  |  |
| LA size, mm | 32.2 ± 5.4 | 32.8 ± 3.8 | 39 | 0.44 |
| TAPSE, mm | 17.3 ± 4.5 | 19.5 ± 3.9 | 30 | 0.016 |
| RAA, end-systolic, cm<sup>2</sup> | 16.2 (14.1, 20.3) | 17.0 (14.0, 21.8) | 21 | 0.73 |
| RVA, end-diastolic, cm<sup>2</sup> | 25.1 (16.3, 34.9) | 28.5 (19.9, 37.7) | 20 | 0.018 |
| *Others* |  |  |  |  |
| SaO<sub>2</sub>, % | 89.3 ± 4.9 | 88.1 ± 5.9 | 38 | 0.38 |
| NT-proBNP, pg/ml | 738.0 (203.0, 2678.0) | 360.1 (145.3, 1602.5) | 41 | 0.74 |
| Refractory pleural effusion, n (%) | 35 (81.3) | 20 (46.5) | 43 | <0.005 |
| CRP, mg/L | 6.3 (2.5, 29.5) | 4.0 (1.4, 16.1) | 37 | 0.054 |
| D-Dimer, ug/ml | 1.2 ± 0.7 | 1.3 ± 1.0 | 36 | 0.67 |

Abbreviations as in Table 1.

## Discussion

In this study, we focused on the incidence and predictors of ISR in PVS-FM. The salient findings are as follows: 1) the incidence of ISR following stent implantation of PVS-FM is as high as 6.3%, 21.4%, and 39.2% at 3-, 6-, and 12-month follow-ups, respectively; 2) RVD is an independent factor for ISR, and the stenosis of the Cor-PA largely affects the occurrence of restenosis.

### In-stent restenosis and associated factors in PV intervention of PVS-FM

Previously, Albers et al.^[4]^ reported a restenosis rate of 7/16 (44%) patients with PVS-FM during a median 115-month follow-up. Similarly, the Mayo Clinic experience described a restenosis rate of up to 4/8 (50%) patients with PVS-FM after the intervention.^[3]^ However, the incidence of restenosis in the above studies might not be accurate and underestimated for the following reasons: 1) the sample size of the above studies was small; 2) three patients were detected by routine CT angiography without associated symptoms, and 4 patients had symptomatic restenosis in the study by Albers et al.^[4]^ The overall median time of 115 months was the time to the symptom of restenosis. 3) In the study from the Mayo Clinic, 2 of the remaining 4 patients died shortly after the procedure.^[3]^ In our study, CT angiography was routinely performed to surveil ISR in 134 PVs of 65 patients with PVS-FM. A total of 61/134 (45.5%) PVs and 42/65 (64.6%) patients had ISR during a median of 6.6 months of follow-up. The cumulative ISR was 6.3%, 21.4%, and 39.2% at the 3-, 6-, and 12-month follow-ups, respectively. Accordingly, this study confirms the high ISR rate with more detailed and accurate information in a larger cohort of PVS-FM patients. A high restenosis rate was also reported in PVS with other etiologies, including PVI. In PVI-PVS, the rate of restenosis is between 19%-39% after a median follow-up period of 6.0-55.2 months.^[16–20]^ Recently, a meta-analysis depicted that the total restenosis rate was 54% in BA and 22.3% in stenting of PVS with different etiologies during a median follow-up time of 13-69 months.^[21]^ Hence, ISR in PVS-FM is higher than that in PVI-PVS. Furthermore, the factors associated with ISR were analyzed by a multiple variables logistic model in this study. We found that RVD and Cor-PA stenosis were independent predictors of ISR following PV intervention in patients with PVS-FM. Previous studies have shown that stent size is associated with restenosis in PVI-PVS. Prieto et al.^[20]^ reported that a stent diameter <10 mm may increase the risk of restenosis. Furthermore, a stent size ≥7 mm was associated with lower restenosis, which was described by Balasubramanian et al.^[22]^ Additionally, some scholars have recommended that a stent diameter >8 mm may serve as the first choice of treatment for PV intervention.^[16]^ Our findings are consistent with previous results to some extent; however, this result is more rational because the choice of stent size is based on the RVD, which was speculated by Prieto et al.^[20]^ who suggested that smaller RVD could increase the rate of restenosis. Meanwhile, the explanation for the higher ISR in PVS-FM than in PVI-PVS is as follows. The involved PVs are often different in diameter. PVI-PVS is always located at the ostia of PV with a larger caliber, while PVS-FM is at the proximal 1^st^ tributary of PV with a smaller caliber. In addition, different pathogeneses could be another attribute. PVI-PVS is intraluminal intimal hyperplasia by physicochemical injury, while PVS-FM is extraluminal by proliferative fibrous tissue compression. PVS-FM might be more easily injured by balloon or stent inflation than PVI-PVS. As expected, Cor-PA stenosis is an exclusive factor associated with ISR, which should be given more attention in PV intervention. Wang et al^[12]^ previously classified FM into 3 types: only the artery involved, only the vein is compressed, and there is both artery and vein narrowing, which should be a mandatory evaluation for an interventional strategy of patients with FM. Overall, the ISR following PV intervention is significantly higher than that following PA intervention, regardless of the etiology of PVS, which may be attributed to the lower pressure in the venous system.^[19]^ A mismatch between the stent and the vessel might increase the occurrence of restenosis.^[23]^ Additionally, it was shown that PVS severity correlated with the severity of the corresponding lung segment in PVS in children.^[24]^

Either over-or underinflation of the balloon or stent may affect restenosis, which has been demonstrated in coronary artery intervention.^[25–27]^ Earlier studies documented some factors of restenosis after percutaneous coronary intervention, including vessel size, maximal balloon pressure, stent type, final diameter stenosis, DM, etc.^[28]^ Endoluminal imaging techniques (e.g., optical coherence tomography, OCT; intravenous ultrasound, IVUS) with existing imaging techniques (e.g., CT angiography, pulmonary venography) may promote standardized management by allowing for intraoperative guidance of stent deployment and postoperative assessment of luminal changes (e.g., thrombosis, intimal hyperplasia) in coronary artery intervention;^[29]^ however, they are limited in PV intervention. Drug-eluting stents have been widely used in coronary artery disease to prevent restenosis.^[23]^ Moreover, Masaki and his team revealed that rapamycin-eluting films could suppress pulmonary vein obstruction progression,^[30]^ which is promising for ISR in PV intervention. In other words, further study is necessary.

### Safety and efficacy of PV intervention of the PVS-FM

A previous study in 8 patients with PVS-FM demonstrated that the incidence of peri-procedure complications and mortality are as high as 3/8 (37.5%) and 3/8 (37.5%), respectively.^[3]^ Another study showed that overall procedure-related complications in patients with FM, including PA, PV, and SVC intervention, were 15/58 (26%) minor and 6/58 (10%) severe.^[4]^ In the present large cohort study, we found that some patients had a cough (19%), chest tightness (18%) and other discomfort (18%), including palpitations, dizziness, nausea, etc. No major hemoptysis or periproced death occurred. The incidence and severity of complications in this study are different from the previous description, which could be attributed to concomitant conditions, inflation pressure, location of PV lesion, and patient status.

In this study, there were immediate improvements in PV caliber, Pd across lesions, and PVFG after the intervention compared with before the intervention, which further supported the findings in previous small sample-sized studies.^[3,4]^ The PAP evaluated by right heart catheterization decreased in the follow-up period. Notably, the lack of improvement in exercise capacity (6MWD, WHO-FC) but having a trend of increase may be attributed to the fact that some of the patients had a recurrence of symptoms or needed further intervention (nearly 50%) during follow-up. On the other hand, remaining PA stenosis may have a negative influence on the overall efficacy. Hence, further long-term follow-up is needed to analyze the efficacy of PV intervention with the PVS-FM.

### Limitations

There are some limitations in this study. First, most ISRs were evaluated by CT venography, which might underestimate or overestimate the degree of restenosis. Second, the sample size was small, and the data were from a single center, which might unavoidably have some bias, even though it was the largest cohort study to date. Third, the follow-up data were not complete, and some patients were lost to follow-up. Finally, no long-term efficacy was followed up because most patients underwent subsequent PA intervention.

### Conclusions

The ISR is very high after the initial intervention of PVS-FM, which is independently associated with RVD and the stenosis of Cor-PA.

## Acknowledgments

All individuals who contributed to this publication have been included as authors.

## Notes

**Funding sources:** This work was supported by the National Natural Science Foundation of China (82070052) and the Open Project of State Key Laboratory of Respiratory Disease (SKLRD-OP-202301).

### Competing Interest Statement

The authors have declared no competing interest.

### Clinical Trial

ChiCTR2000033153

### Funding Statement

This work was supported by the National Natural Science Foundation of China (82070052) and the Open Project of State Key Laboratory of Respiratory Disease (SKLRD-OP-202301).

